# Pooling of samples for SARS-CoV-2 surveillance using a rapid antigen test

**DOI:** 10.1101/2021.02.09.21250610

**Authors:** Nol Salcedo, Alexander Harmon, Bobby Brooke Herrera

## Abstract

While molecular assays, such as RT-PCR, have been widely used throughout the COVID-19 pandemic, the technique is costly and resource intensive. As a means to reduce costs and increase diagnostic efficiency, pooled testing for RT-PCR has been implemented. However, pooling samples for antigen testing has not been evaluated. We propose a pooling strategy for antigen testing that would significantly expand SARS-CoV-2 surveillance, especially for low-to-middle income countries, schools, and workplaces. Our data demonstrate that combining of up to 20 nasal swab specimens per pool can expand surveillance with antigen tests, even if a pool contains only one positive sample.

## Introduction

With ongoing worldwide transmission of severe acute respiratory syndrome coronavirus 2 (SARS-CoV-2) and emergent variants, laboratory testing has played an important role in detecting the virus in symptomatic and asymptomatic individuals. Molecular assays, such as reverse-transcription polymerase chain reaction (RT-PCR), are accurate but costly and resource and supply chain intensive, challenging large scale surveillance efforts, especially for low-to-middle income countries. The financial and logistical barriers associated with RT-PCR restricts the ability to rapidly identify and quarantine infected individuals and hinder our ability to estimate community prevalence and epidemic trajectory. These limitations underscore the need for novel and dynamic approaches to individual diagnostics, community surveillance, and epidemic control.

Pooling subsamples and processing them in groups by RT-PCR has been proposed as a testing strategy to reduce costs and increase efficiency. If a pool tests negative by RT-PCR, then all of the constituent samples are assumed negative. If a pool tests positive by RT-PCR, then the constituent samples are putatively positive and must be re-tested individually or in mini pools. A recent study demonstrated that a range of positive pools could be identified, even if the Ct value of a single sample was up to 34. Simple RT-PCR pooling designs can also be used to assess prevalence without individual specimen identification, by using quantitative viral load measurements in each positive pool, where the viral load measurement from a pool is proportional to the sum of the diluted viral loads from each positive sample in the pool.

Public health surveillance tools, such as rapid antigen tests, have also been advocated for since early in the pandemic to help control SARS-CoV-2 spread. Rapid antigen tests are optimized to detect the presence of SARS-CoV-2 proteins in individuals during the acute, infectious phase of COVID-19, and can be self-administered or performed at the point-of-care, leading to faster results from the time of sampling and more frequent testing. However, pooled testing using rapid antigen tests, has not been evaluated. In this study, we examined a pooling strategy for public health surveillance of SARS-CoV-2 using a rapid antigen test.

## Methods

### Clinical Dilution Panel

The study included a clinical dilution panel provided by the non-profit PATH (www.path.org). The panel was prepared from human nasal swab eluate discards in phosphate-buffered saline. Swab eluates positive for SARS-CoV-2 by RT-PCR from multiple individuals were combined and diluted into nasal swab eluate pools from individuals testing negative for SARS-CoV-2 by RT-PCR. Dilutions were then aliquoted and frozen at -80°C.

The primary studies under which the samples were collected received ethical clearance from PATH Institutional Review Board (IRB) (approval number 00004244). All excess samples and corresponding data were banked and de-identified prior to analysis. This study received an exemption determination from the PATH IRB.

### Quantitative Reverse-Transcriptase Polymerase Chain Reaction

Aliquots from the clinical dilution panel were thawed and 200 μl was used for extraction with the QIAamp Viral RNA Mini Kit (Qiagen, Germany). Nucleic acids were eluted in 50 μl and 10 μl were used for qRT-PCR using the CDC’s 2019-nCoV Real-Time RT-PCR N1 assay on the QuantStudio 5 Real-Time PCR System (Applied Biosystems, Foster City, CA). Viral loads were determined by measuring the concentration of the RNA using a previously qualified standard to determine genomic RNA levels.

### Sample Pooling

For the 20-fold pooled sample testing, 50 μl from a single RT-PCR confirmed positive nasal swab specimen dilution was mixed with 50 μl from nineteen RT-PCR confirmed negative nasal swab specimens. The total volume of each pool was 1 ml. For the non-pooled testing, the RT-PCR confirmed nasal swab specimen dilutions were not mixed with any negative nasal swab specimens, and were tested directly on the rapid antigen test. For pooled and single sample testing, 100 μl of the sample was applied to the COVID-19 rapid antigen test and allowed to react for 15 minutes before test result image capture.

### COVID-19 Antigen Test

The rapid antigen test (E25Bio, Inc., Cambridge, MA) contains a monoclonal antibody and a nanoparticle conjugate that detects the nucleocapsid protein of SARS-CoV-2. Interaction of the immobilized Test and Control line antibodies with antigen and the nanoparticle conjugate produces visible bands, indicating whether the test is positive or negative. The lot used in this study was validated using recombinant SARS-CoV-2 nucleocapsid protein (SinoBiological, China) from 0.1 ng/ml to 500 ng/ml in a total volume of Solution Buffer (E25Bio, Inc., Cambridge, MA) of 100 μl (data not shown).

### Image Analysis

Rapid antigen test result images were captured via the iPhone E25Bio Passport application (currently only available via TestFlight) and analyzed using image processing software, Image J (NIH), to machine-read and quantify test results. The average pixel intensity was quantified at the Test line, Control line, and background areas. The background-adjusted Test line signal was then normalized to the background-subtracted Control line and expressed as an arbitrary unit (A.U.).

## Results

To analyze the effect of sample pooling on the analytical sensitivity of a rapid antigen test, we compared antigen detectability of single RT-PCR confirmed nasal swab specimens with varying Ct values versus when pooled into 19 RT-PCR confirmed negative nasal swab specimens. Antigen detectability was determined by applying to a rapid antigen test 100 μl of a single RT-PCR positive nasal swab specimen or 100 μl of a pooled sample. Each pooled sample consisted of 50 μl of a RT-PCR positive nasal swab dilution specimen and a 950 μl negative nasal swab mixture (50 μl from 19 RT-PCR confirmed negative nasal swab specimens).

Our results show that the rapid antigen test detected positive pools between Ct values 18.3 (VL: 7.6E7) and 30.1 (VL: 4.8E4), compared to 18.3 and 31.9 (VL: 3.5E4) for single nasal swab specimens (Fig. 1). These data suggest that combining of up to 20 samples per pool can expand surveillance with rapid antigen tests, even if a pool contains only one positive sample. Only in the case of a positive pool test result is additional rapid antigen testing and/or RT-PCR of individual samples or smaller pools required.

**Fig. 1.**
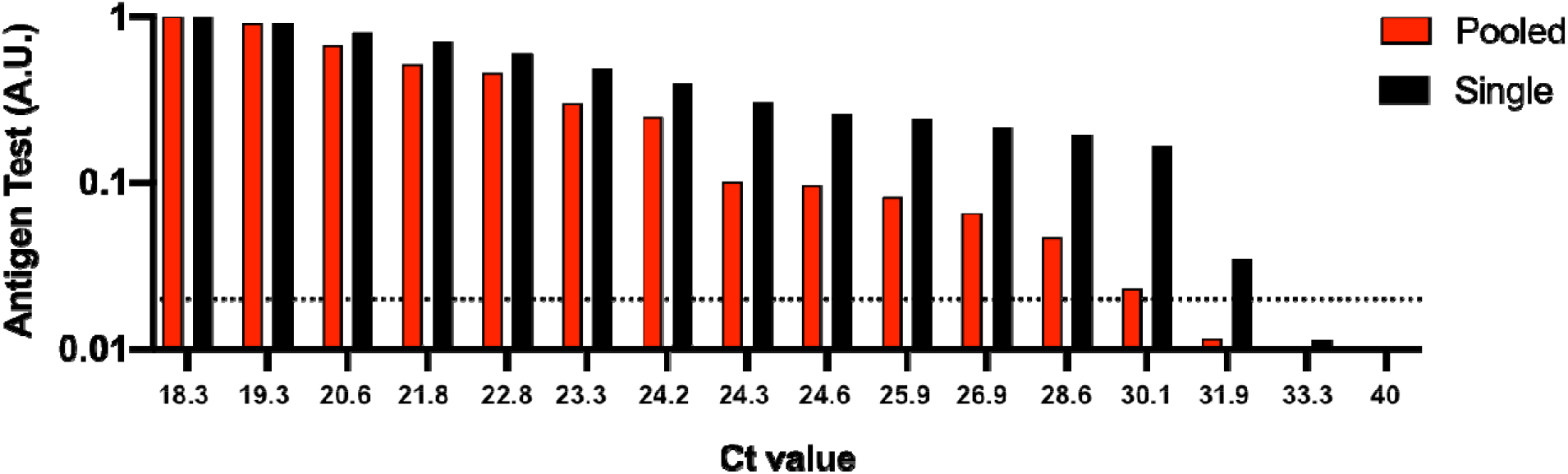
Analytical sensitivity of a rapid antigen test using single nasal swab specimens versus when pooled. 50 μl of the positive nasal swab specimens with Ct values ranging between 18.3 and 40 were spiked into 50 μl of RT-PCR confirmed negative nasal swab specimens from 19 individuals. 100 μl of the 1 ml pooled samples or 100 μl of the single positive nasal swab specimens were applied to the rapid antigen test and allowed to react for 15 minutes before test results were image captured. To determine relative nucleocapsid antigen levels detected by the antigen test, images were analyzed using image processing software and reported as an arbitrary unit (A.U.). Ct, cycle threshold. Horizontal dashed line, limit of detection for the rapid antigen test.

## Discussion

We propose a pooling strategy for antigen testing that is easy to implement and can further expand SARS-CoV-2 surveillance. In general, for single sample testing, a nasal swab is collected from an individual, resuspended into 0.5-1 ml solution (e.g. viral transport media, saline solution, antigen test kit buffer solution, etc.), and 100 μl of the nasal swab specimen is then applied to a rapid antigen test. However, performing individual antigen testing on a single nasal swab specimen can present further cost and logistical limitations when the frequency of antigen testing is increased, especially for wide-scale surveillance. For pooled sample testing, nasal swabs are collected from up to 20 individuals, each of the nasal swabs are then resuspended into 0.3-0.5 ml solution, and 50 μl from each nasal swab specimen is pooled into a mixture before applying 100 μl to a rapid antigen test (Fig. 2). If a pool tests negative by an antigen test, then all of the constituent nasal swab specimens are assumed negative. If a pool tests positive by the antigen test, then the constituent nasal swab specimens are putatively positive and should be re-tested in mini pools. If a mini pool remains positive, then antigen testing on individual nasal swab specimens can be performed.

**Fig. 2.**
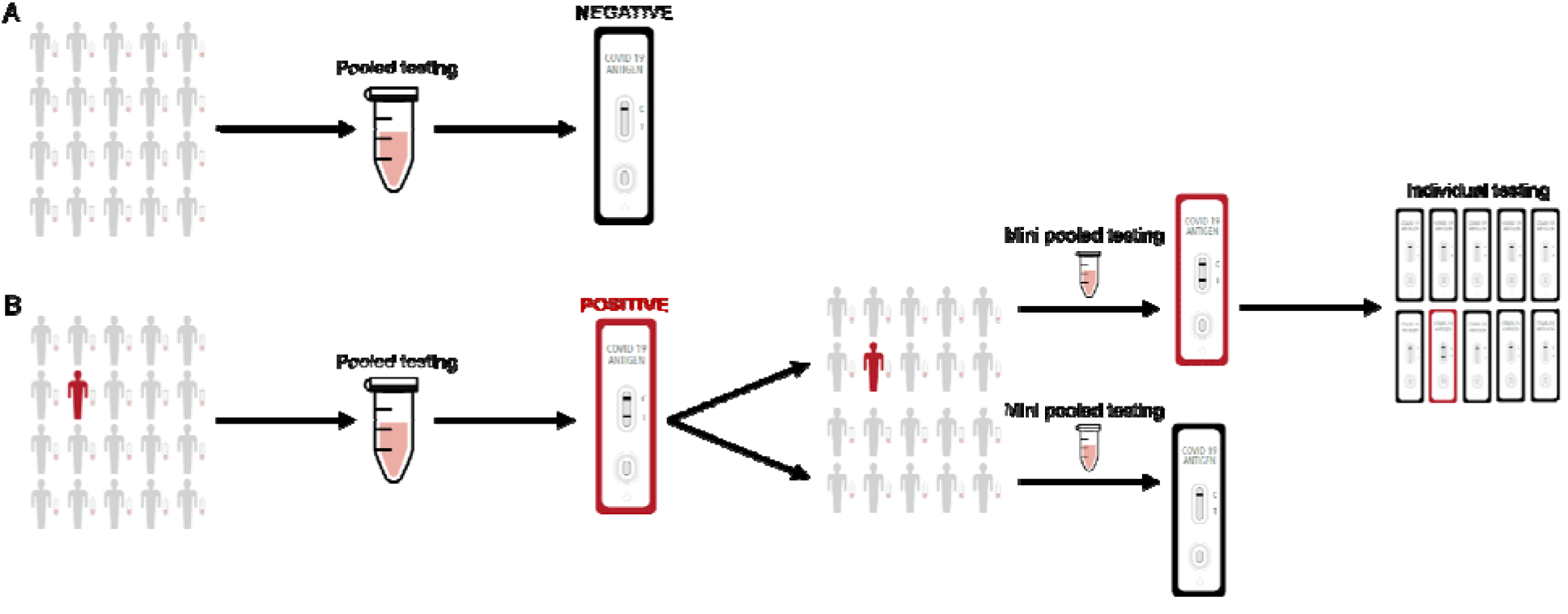
Sample pooling strategy for antigen testing. A) Nasal swab specimens are collected, where 50 μl from each individual is pooled into a single mixture and 100 μl is then applied to the rapid test. If a pool tests negative by the antigen test, then all of the constituent nasal swab specimens are assumed negative. B) If a pool tests positive by the antigen test, then the constituent nasal swab specimens are putatively positive and should be re-tested in mini pools. If a mini pool remains positive, then antigen testing on individual nasal swab specimens can be performed.

Pooled testing using rapid antigen tests is a cost-effective and public health conscious approach to expanding SARS-CoV-2 surveillance, thus further enabling societal reopening, especially in instances where capacity and resources are constrained. Our results show that the rapid antigen test can detect SARS-CoV-2 positive nasal swab specimens up to Ct value 30.1, even when the positive specimen is combined with 19 negative nasal swab specimens.

While our experiments suggest that a pooling strategy for antigen testing may be beneficial for SARS-CoV-2 surveillance and identification of individuals during the infectious phase of COVID-19, there are limitations. Specimens collected and tested beyond the acute infection phase (7 days post-infection) will likely escape detection by a rapid antigen test, regardless if the specimen is pooled or non-pooled prior to testing. Enhancing efficiency at the expense of sensitivity must be appropriately considered depending on the purpose of testing and resources available. A number of modeling and clinical studies have shown that increasing the frequency of antigen testing to a minimum of two times per week can increase the performance metrics of an antigen test. Furthermore, pooling adds complexity because samples must be archived, or individuals would need to undergo additional swabbing for potential re-testing.

We have shown that a simple design for antigen testing using a sample pooling strategy is straightforward and can enhance SARS-CoV-2 surveillance. Future studies will be important to determine whether pooling strategies can be applied to COVID-19 antibody detection in sera using rapid tests or for antigen detection from other microorganisms. While there are other costs and logistical limitations associated with pooling that we do not consider here, our study highlights the use of antigen tests as a public health tool to help combat the COVID-19 pandemic.

## Data Availability

All data is available within the manuscript; should you require additional data please contact the corresponding author.

## References

Cleary, Brian, James A. Hay, Brendan Blumenstiel, Maegan Harden, Michelle Cipicchio, Jon Bezney, Brooke Simonton, et al. “Using Viral Load and Epidemic Dynamics to Optimize Pooled Testing in Resource Constrained Settings.” medRxiv (2021): 2020.05.01.20086801. https://doi.org/10.1101/2020.05.01.20086801. https://www.medrxiv.org/content/medrxiv/early/2021/01/15/2020.05.01.20086801.full.pdf.

Larremore, D. B., B. Wilder, E. Lester, S. Shehata, J. M. Burke, J. A. Hay, M. Tambe, M. J. Mina, and R. Parker. “Test Sensitivity Is Secondary to Frequency and Turnaround Time for Covid-19 Screening.” Sci Adv (Nov 20 2020). https://doi.org/10.1126/sciadv.abd5393. https://www.ncbi.nlm.nih.gov/pubmed/33219112.

Lauring, A. S., and E. B. Hodcroft. “Genetic Variants of Sars-Cov-2-What Do They Mean?”. JAMA 325, no. 6 (Feb 9 2021): 529-31. https://doi.org/10.1001/jama.2020.27124. https://www.ncbi.nlm.nih.gov/pubmed/33404586.

Lohse, S., T. Pfuhl, B. Berko-Gottel, J. Rissland, T. Geissler, B. Gartner, S. L. Becker, S. Schneitler, and S. Smola. “Pooling of Samples for Testing for Sars-Cov-2 in Asymptomatic People.” Lancet Infect Dis 20, no. 11 (Nov 2020): 1231–32. https://doi.org/10.1016/S1473-3099(20)30362-5. https://www.ncbi.nlm.nih.gov/pubmed/32530425.

Mahmoud, S. A., E. Ibrahim, B. Thakre, J. G. Teddy, P. Raheja, S. Ganesan, and W. A. Zaher. “Evaluation of Pooling of Samples for Testing Sars-Cov-2 for Mass Screening of Covid-19.” BMC Infect Dis 21, no. 1 (Apr 17 2021): 360. https://doi.org/10.1186/s12879-021-06061-3. https://www.ncbi.nlm.nih.gov/pubmed/33865325.

Nash, Beatrice, Anthony Badea, Ankita Reddy, Miguel Bosch, Nol Salcedo, Adam R. Gomez, Alice Versiani, et al. “Validating and Modeling the Impact of High-Frequency Rapid Antigen Screening on Covid-19 Spread and Outcomes.” medRxiv (2021): 2020.09.01.20184713. https://doi.org/10.1101/2020.09.01.20184713. https://www.medrxiv.org/content/medrxiv/early/2021/01/30/2020.09.01.20184713.full.pdf.

Peeling, R. W., P. L. Olliaro, D. I. Boeras, and N. Fongwen. “Scaling up Covid-19 Rapid Antigen Tests: Promises and Challenges.” Lancet Infect Dis (Feb 23 2021). https://doi.org/10.1016/S1473-3099(21)00048-7. https://www.ncbi.nlm.nih.gov/pubmed/33636148.

Tromberg, B. J., T. A. Schwetz, E. J. Perez-Stable, R. J. Hodes, R. P. Woychik, R. A. Bright, R. L. Fleurence, and F. S. Collins. “Rapid Scaling up of Covid-19 Diagnostic Testing in the United States - the Nih Radx Initiative.” N Engl J Med 383, no. 11 (Sep 10 2020): 1071–77. https://doi.org/10.1056/NEJMsr2022263. https://www.ncbi.nlm.nih.gov/pubmed/32706958.

Wiersinga, W. J., A. Rhodes, A. C. Cheng, S. J. Peacock, and H. C. Prescott. “Pathophysiology, Transmission, Diagnosis, and Treatment of Coronavirus Disease 2019 (Covid-19): A Review.” JAMA 324, no. 8 (Aug 25 2020): 782–93. https://doi.org/10.1001/jama.2020.12839. https://www.ncbi.nlm.nih.gov/pubmed/32648899.

